# DNA Methylation of Neurodevelopmental, Stress-Response, Reward, and Opioid Pathway Genes: Sex Differences in Infants with Neonatal Opioid Exposure

**DOI:** 10.64898/2026.09.28.26364198

**Authors:** Angélica Aragón Vásquez, Marie Camerota, Mara G. Coyle, Elisabeth D. Conradt, Barry M. Lester

## Abstract

**Objective:** To investigate sex-specific associations between DNA methylation (DNAm) of neurodevelopmental, stress-response, and dopaminergic pathway genes and need for pharmacological treatment for Neonatal Opioid Withdrawal Syndrome (NOWS) among opioid-exposed newborns.

**Study Design:** Buccal swabs were collected at birth from 181 infants in the multi-site Child and Family Study (CAFS). DNAm was measured via pyrosequencing at CpG sites within promoter regions of neurodevelopmental (*BDNF*), stress-response (*AVP, NR3C1, OXTR, FKBP5, SLC6A4*), dopaminergic (*DRD2, DRD4*), and opioid signaling (*OPRM1*) genes. Need for pharmacological treatment for NOWS was obtained from medical records. We estimated robust linear regression models to investigate sex-specific associations of DNAm and need for NOWS treatment. Estimated marginal means quantified sex-specific DNAm differences by treatment status.

**Results:** After correction for multiple testing, five CpG sites within *DRD4* were significantly associated with need for pharmacological treatment, with four sites showing higher DNAm in treated infants and one showing lower DNAm. Significant sex-specific associations were identified within *DRD4* and *AVP*.

**Conclusions:** Epigenetic variation across multiple biological pathways contributes to variability in NOWS severity among infants with prenatal opioid exposure. The relationship between epigenetic variation and need for NOWS treatment may vary depending on infant sex.

## Introduction

The opioid epidemic has profoundly impacted maternal and infant health in the United States, with prenatal opioid exposure recognized as a major public health concern. Maternal opioid-related diagnoses at delivery increased by 131% between 2010 and 2017, while diagnoses of Neonatal Opioid Withdrawal Syndrome (NOWS) increased by 82%, from 4.0 to 7.3 per 1,000 hospital births.^1^ Critically, not all infants with prenatal opioid exposure develop NOWS, and the biological mechanisms determining which infants will experience NOWS are unknown.^2^ Infant sex may represent one biological contributor, as male infants tend to exhibit more severe NOWS symptoms and receive pharmacological treatment more frequently than females.^3, 4^ Sex-specific differences in molecular responses to prenatal opioid exposure have also been observed, including higher *DRD2* expression in male newborns with NOWS relative to females^5^, suggesting that sex-specific biological mechanisms may contribute to variability in NOWS severity.

Epigenetic variation represents a pathway through which prenatal opioid exposure may influence NOWS severity. Most epigenetic studies of NOWS have focused narrowly on *OPRM1*, which encodes the *mu*-opioid receptor, where increased DNA methylation (DNAm) has been associated with greater NOWS severity, including need for pharmacological treatment.^6, 7^ NOWS reflects dysregulation across stress physiology, neurobehavioral regulation, and dopaminergic reward systems^8, 9^, yet DNAm of genes involved in these broader pathways has not been systematically examined in relation to NOWS. Moreover, despite documented differences in NOWS for males and females^3, 4^, there has yet to be comprehensive investigation of sex-specific associations between DNAm and NOWS severity.

The objective of the present study was to investigate associations between DNAm in regulatory regions of neurodevelopmental (*BDNF*), stress-response (*AVP, NR3C1, OXTR, FKBP5, SLC6A4*), dopaminergic (*DRD2, DRD4*), and opioid signaling (*OPRM1*) pathway genes and need for pharmacological treatment among newborns exposed prenatally to opioids. We also aimed to test whether these associations differed by infant sex. We hypothesized that DNAm within regulatory regions of these genes would differ between infants who required pharmacological treatment for NOWS and those who did not, and that these associations would vary by infant sex.

## Methods

### Participants

Participants in the Child and Family Study (CAFS) were part of a prospective, multisite investigation aimed at identifying novel clinical indicators of NOWS. Recruitment of 235 mother-infant dyads with prenatal exposure to opioids took place at Women and Infants Hospital of Rhode Island (WIH) and the University of Utah Hospital between 2019 and 2025. Mothers were approached for consent if prenatal opioid exposure was identified either during pregnancy or delivery as indicated by (1) maternal medical record, (2) a positive maternal urine toxicology during pregnancy or at time of hospital admission, (3) a positive infant toxicology screen via umbilical cord or urine or (4) treatment for opioid use disorder. Exclusion criteria included congenital anomalies, genetic syndromes, metabolic disturbances, sepsis, asphyxia, seizures, respiratory failure, a gestational age at birth of < 33 weeks, inability to take oral medication, inability of the caregiver to provide informed consent, and inability to tolerate a newborn neurobehavioral exam. The Institutional Review Board approved procedures for each study site, and written consent was obtained from all participants.

### Need for pharmacological treatment for NOWS

Information regarding NOWS symptoms and need for pharmacological treatment was obtained from the infant’s medical records. Assessment of withdrawal severity and criteria for initiating pharmacological treatment followed site-specific clinical protocols. At WIH, NOWS symptoms were evaluated using the Finnegan Neonatal Abstinence Syndrome Scoring Tool (FNAST). Pharmacological treatment was initiated following either three consecutive FNAST scores greater than 7 or two consecutive scores greater than 11. At the University of Utah, withdrawal symptoms were assessed using the Neonatal Withdrawal Inventory (NWI) and the Eat, Sleep, Console (ESC) approach. The NWI is an empirically derived eight-item instrument adapted from the FNAST, with treatment initiated when an infant received one or more scores of 8 or higher. The ESC method evaluates functional indicators of withdrawal, including the infant’s ability to feed appropriately for age, sleep undisturbed for at least one hour between care periods, and be consoled within 10 minutes; failure to meet any of these criteria could prompt initiation of pharmacological treatment. Pharmacological treatment for NOWS at both sites primarily consisted of oral morphine, with phenobarbital administered as additional therapy for infants who exceeded a predetermined morphine dose threshold.

### DNA methylation

DNA was extracted from buccal swabs collected shortly after birth, prior to initiation of pharmacological treatment (*M*_age_ = 1.5 days of life, *SD*_age_ = 0.96 days, range = 0 to 6 days). Quantitative DNAm was analyzed as described previously.^10^ In summary, QIAamp DNA Mini Kits (Qiagen, Inc.) were used to extract genomic DNA by following the manufacturer’s protocols. Then, pyrosequencing (Qiagen Inc.) was used on the purified DNA to carry out quantitative methylation analysis. The EZ DNA Methylation Kit (Zymo Research) was used to bisulfite-modify the DNA samples (1μg). The PCR amplified products from the bisulfite-modified DNA were pyrosequenced on a Qiagen Q48 pyrosequencing platform using assay reagents from EpigenDx for CpG sites within promoter regions of neurodevelopmental (*BDNF*), stress-response (*AVP, NR3C1, OXTR, FKBP5, SLC6A4*), dopaminergic (*DRD2, DRD4*), and opioid signaling (*OPRM1*) pathway genes. The performance characteristics of the assay were evaluated using a dilution series of fully methylated reference DNA into fully unmethylated reference DNA. DNAm values are reported as percent methylation at each CpG site. Standard assay quality control procedures were applied, and CpG sites with unreliable signals were excluded prior to analysis.

### Statistical Analysis

DNAm of individual CpG sites within genes of interest were analyzed to examine differences between infants who required pharmacological treatment and those who did not. Analyses were conducted in a stepwise manner. Robust linear regression and non-parametric tests were selected due to the non-normal distribution commonly observed in DNAm data. First, unadjusted group differences between infants who required pharmacological treatment and those who did not were assessed using Wilcoxon rank-sum tests. Robust linear regression models (RLMs) were then used to examine associations between DNAm at each CpG site and subsequent need for pharmacological treatment, controlling for infant sex and study site. These covariates were chosen due to known sex differences in neurodevelopment and differences in clinical management of NOWS across sites. To evaluate whether associations between DNAm and need for pharmacological treatment differed by sex, additional RLMs were fitted that included a sex-by-treatment interaction term, with study site retained as a covariate. To correct for multiple comparisons, *p*-values from the regression models were adjusted using the false discovery rate (FDR) within each gene using the Benjamini–Hochberg method. Statistical significance was defined as FDR-adjusted *p*≤.10. For CpG sites with a significant Sex x Treatment interaction term, estimated marginal means (EMMs) were calculated from the robust linear regression models to quantify adjusted DNAm levels by treatment status and infant sex. All analyses were conducted in R (version 4.5.2). RLMs were fit using the MASS package, and estimated marginal means were calculated using the emmeans package. Analyses were conducted using complete case data.

## Results

Of the 235 enrolled mother–infant dyads, 181 had DNAm data and were included in the analyses. There were no significant differences in maternal and infant characteristics between included and excluded dyads except for maternal bipolar disorder diagnosis, which was more common in the included dyads (26.4% vs 9.4%, *p* = 0.02; **Supplemental Table 1**).

Overall, 76 infants (42%) required pharmacological treatment for NOWS, and 89 infants (49%) were male (**Table 1**). Need for treatment did not vary by infant sex (42.7% in males, 41.3% in females, *p* = 0.88). No significant differences by treatment status or sex were observed for sociodemographic, substance use, polypharmacy or medical variables (all *p* > .05). One exception was that mothers of infants who required pharmacological treatment were more likely to have a diagnosis of bipolar disorder compared to mothers of infants who did not require pharmacological treatment (36% vs. 19%, *p* = .02).

**Table 1:** Maternal and Infant Characteristics by Infant Pharmacological Treatment Group and Sex.

|  | **Overall**<br>**N = 181** | **Treatment**<br>**N = 76** | **No**<br>**Treatment**<br>**N = 105** |  | **Male**<br>**N = 89** | **Female**<br>**N = 92** |  |
| --- | --- | --- | --- | --- | --- | --- | --- |
|  | *M (SD) or*<br>*N (%)* | *M (SD) or N*<br>*(%)* | *M (SD) or N*<br>*(%)* | **p-**<br>**value** | *M (SD)*<br>*or N (%)* | *M (SD)*<br>*or N (%)* | **p-**<br>**value** |
| **Maternal characteristics** |  |  |  |  |  |  |  |
| *Less than high school degree* | 45 (25%) | 15 (20%) | 30 (30%) | 0.20 | 22 (26%) | 23 (26%) | 1.00 |
| *Adequate prenatal care* | 146 (81%) | 63 (83%) | 83 (79%) | 0.74 | 73 (82%) | 73 (80%) | 0.91 |
| *No partner* | 77 (43%) | 32 (43%) | 45 (43%) | 1.00 | 39 (44%) | 38 (42%) | 0.85 |
| *Hypertension* | 39 (22%) | 19 (25%) | 20 (19%) | 0.41 | 17 (19%) | 22 (24%) | 0.57 |
| *Residential Treatment* | 40 (22%) | 15 (20%) | 25 (24%) | 0.64 | 19 (21%) | 21 (23%) | 0.95 |
| *Inpatient detox during pregnancy* | 34 (19%) | 13 (17%) | 21 (21%) | 0.67 | 14 (17%) | 20 (21%) | 0.42 |
| *Bipolar* | 47 (26%) | 27 (36%) | 20 (19%) | 0.02* | 20 (23%) | 27 (30%) | 0.40 |
| *PTSD* | 66 (37%) | 32 (43%) | 34 (33%) | 0.25 | 36 (37%) | 30 (33%) | 0.31 |
| *Hepatitis C* | 60 (35%) | 28 (39%) | 32 (31%) | 0.35 | 27 (32%) | 33 (37%) | 0.60 |
| *Polypharmacy ($\geq 2$ classes)* | 167 (92%) | 71 (93%) | 96 (91%) | 0.831 | 85 (96%) | 82 (89%) | 0.185 |
| **Infant characteristics** |  |  |  |  |  |  |  |
| *Sex (Male)* | 89 (49%) | 38 (50%) | 51 (49%) | 0.88 | -- | -- | -- |
| *Gestational age (weeks)* | 38.29 (1.7) | 38.42 (1.8) | 38.2 (1.7) | 0.41 | 38.23 (1.7) | 38.35 (1.8) | 0.65 |
| *Birth weight (grams)* | 3006.41 (551.1) | 2984.47 (567.6) | 3022.29 (541) | 0.65 | 3077.07 (536.1) | 2938.05 (559.6) | 0.09 |
| *Head circumference (cm)* | 33.61 (1.9) | 33.36 (1.8) | 33.8 (1.9) | 0.12 | 33.9 (1.9) | 33.34 (1.8) | 0.06 |
| *Birth length (cm)* | 48.45 (3.0) | 48.28 (3.3) | 48.59 (2.9) | 0.501 | 48.86 (3.1) | 48.05 (2.9) | 0.08 |
| *Social Services involvement* | 85 (47%) | 46 (61%) | 39 (38%) | 0.004 * | 38 (43%) | 47 (52%) | 0.29 |
| *Breastfeeding* | 108 (60%) | 35 (46%) | 73 (70%) | 0.003 * | 50 (56%) | 58 (63%) | 0.43 |
| *Maximum Finnegan Score* | 10.3 (3.0) | 12.34 (2.2) | 7.8 (1.8) | <0.001 * | 10.31 (2.9) | 10.29 (3.1) | 0.97 |

Note: Percentages are calculated among participants with available data for each variable;
denominators may therefore vary across measures.

Infant gestational age, birth weight, head circumference, and birth length did not differ by treatment status (all *p* > .05). However, infants who required pharmacological treatment for NOWS were more likely to have Social Services involvement (61% vs. 38%, *p* = .004) and were less likely to be breastfed (46% vs. 70%, *p* = .003). As expected, infants who required pharmacological treatment had higher maximum FNAST scores compared to those who did not (*M* = 12.34 vs. 7.80, *p* < .001).

After FDR adjustment, there was a main effect of DNAm on need for NOWS within the *DRD4* gene. Four CpG sites within the *DRD4* 5′ regulatory region demonstrated higher DNAm in infants who later required pharmacological treatment relative to those that did not (β = 0.47, distance from TSS = −351; β = 0.31, distance from TSS = −338; β = 0.43, distance from TSS = −303; β = 0.87, distance from TSS = −301; **Table 2**), while one site showed significantly lower DNAm in infants who later received pharmacological treatment (β = −0.27, distance from TSS = −333; **Table 2**). **Figure 1** illustrates the distribution of DNAm levels at individual *DRD4* CpG sites by pharmacological treatment status. Results for all CpGs in all genes are shown in **Supplemental Table 2**.

**Table 2:** Summary of Model Results from *DRD4* Promoter Region CpG Sites.

| Distance from Transcription Start Site | Treatment Adjusted M | No Treatment Adjusted M | B Treatment (SE) | Raw *p*-Value | False Discovery Rate *p*-value |
| --- | --- | --- | --- | --- | --- |
| -362 | 6.22 | 5.57 | 0.66 (0.79) | 0.40 | 0.58 |
| -358 | 0.70 | 0.64 | 0.06 (0.13) | 0.63 | 0.68 |
| -351 | 1.34 | 0.86 | 0.47 (0.19) | 0.01* | 0.10* |
| -346 | 4.18 | 3.87 | 0.31 (0.48) | 0.52 | 0.61 |
| -342 | 1.71 | 1.49 | 0.22 (0.23) | 0.34 | 0.56 |
| -338 | 0.87 | 0.56 | 0.31 (0.15) | 0.04* | 0.10* |
| -333 | 0.42 | 0.69 | -0.27 (0.12) | 0.03* | 0.10* |
| -321 | 1.80 | 1.79 | 0.01 (0.30) | 0.97 | 0.97 |
| -315 | 1.31 | 1.07 | 0.23 (0.24) | 0.34 | 0.56 |
| -312 | 0.51 | 0.43 | 0.07 (0.11) | 0.51 | 0.61 |
| -305 | 2.02 | 1.60 | 0.42 (0.28) | 0.13 | 0.27 |
| -303 | 1.49 | 1.06 | 0.43 (0.19) | 0.02* | 0.10* |
| -301 | 3.43 | 2.56 | 0.87 (0.39) | 0.03* | 0.10* |

*Note:* Treatment and no-treatment values represent adjusted means (emms) from robust linear regression models controlling for infant sex and study site. $\beta$ coefficients represent adjusted treatment effects from the same models. \*Indicates raw $p < 0.05$ or FDR-adjusted $p \leq 0.10$ .

**Figure 1:**
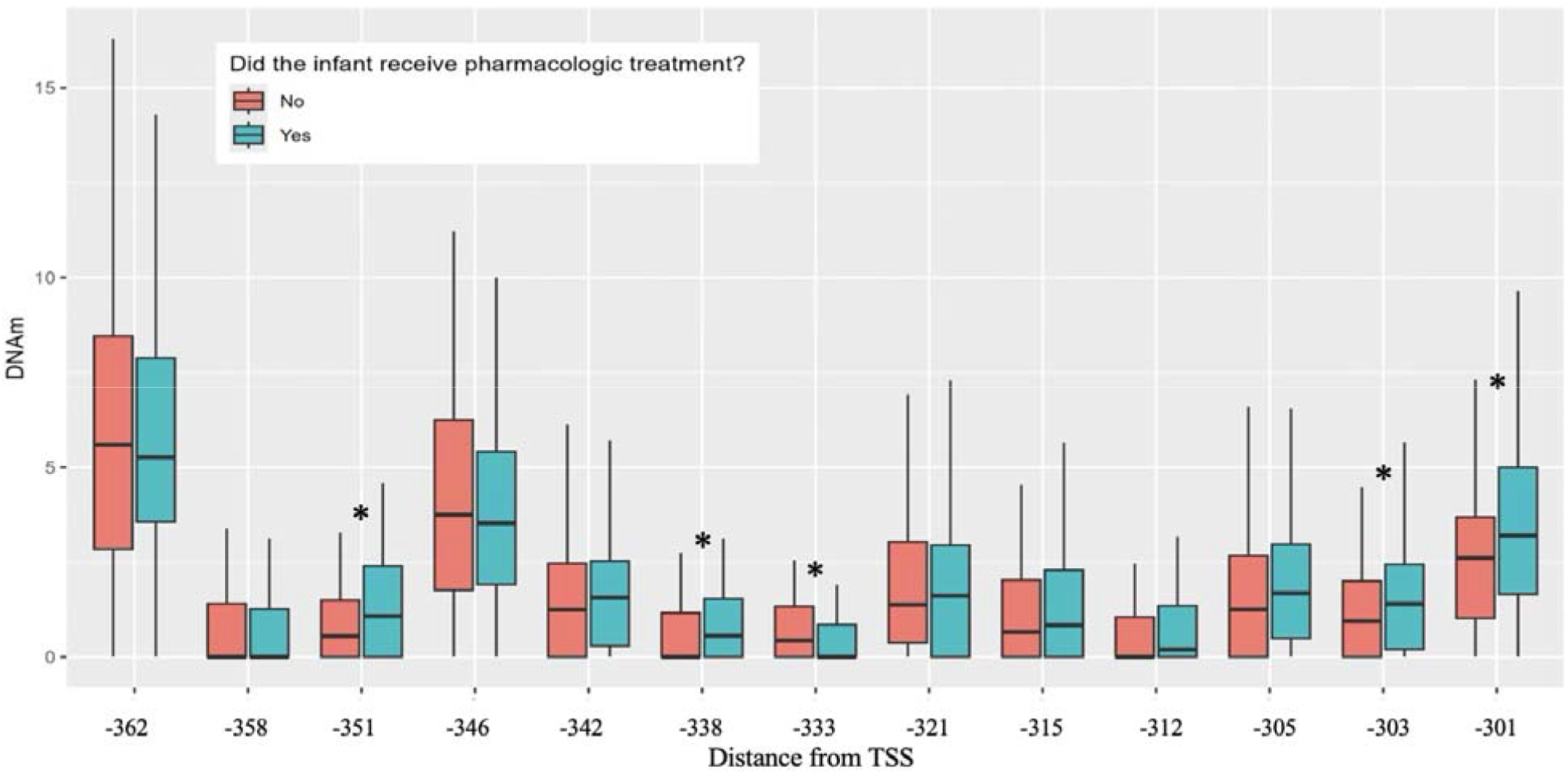
DNA methylation levels at *DRD4* CpG sites by pharmacological treatment status. CpG sites are identified by distance in base pairs from the transcription start site (TSS). Five CpG sites showed significant differences in DNAm by treatment status after false discovery rate (FDR) adjustment. *\**Indicates FDR adjusted *p* ≤ .*10*

Significant Sex × Treatment interactions were identified at two CpG sites after FDR adjustment: one within *DRD4* (β = 3.65, *p* = 0.04) and one within *AVP* (β = 2.98, *p* = 0.04). For the *DRD4* CpG located −362 from TSS, we observed higher DNAm among male infants who required pharmacological treatment for NOWS relative to those that did not (M_treated_ = 8.38, M_untreated_ = 5.75, *p* < .05; **Figure 2A**). There was no difference in DNAm by treatment status for female infants at this site (M_treated_ = 5.90, M_untreated_ = 6.92, *p* = 0.22; **Figure 2A**).

**Figure 2:**
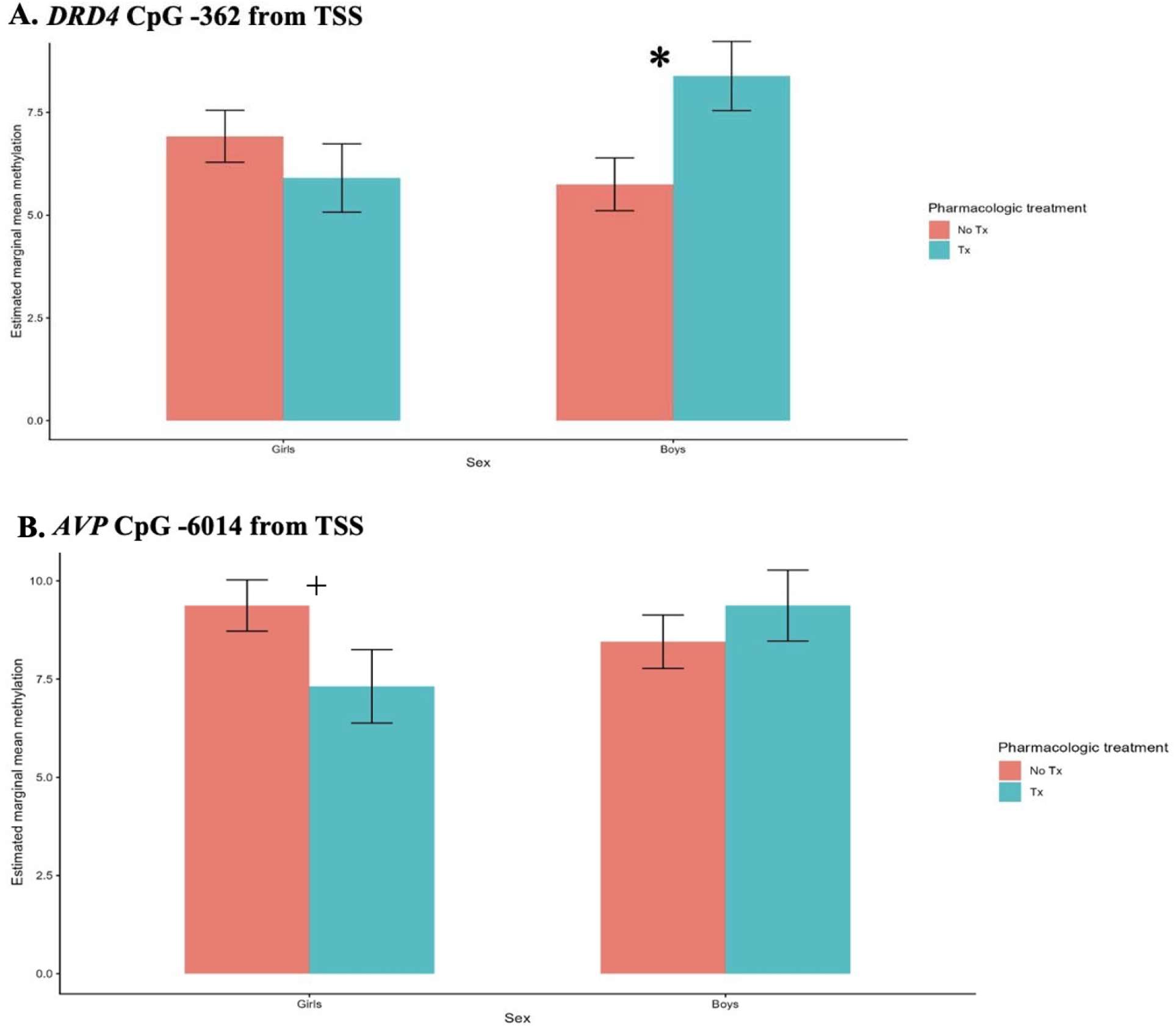
Estimated marginal means of significant sex × pharmacological treatment interactions at **A**, the *DRD4* CpG site −362 bp from the TSS, and **B**, the *AVP* CpG site −6014 bp from the TSS. *\**Indicates *p* ≤ .05 and + Indicates *p* = .06.

For the *AVP* CpG located −6014 from the TSS, although the interaction effect was significant, comparison of marginal means showed a nonsignificant trend toward lower DNAm among female infants requiring pharmacological treatment compared with those who did not require treatment (M_treated_ = 7.32, M_untreated_ = 9.37, *p* = .06; **Figure 2B**), while no DNAm differences were observed among male infants (M_treated_ = 9.37, M_untreated_ = 8.45, *p* = 0.28; **Figure 2B**).

## Discussion

We investigated associations between DNAm in regulatory regions of genes involved in stress signaling, neurodevelopment, dopaminergic and opioid receptor pathways and need for pharmacological treatment for NOWS among newborns with prenatal opioid exposure, including how these relationships differed by infant sex. We found that differential DNAm of several CpG sites within the regulatory region of *DRD4* was associated with need for pharmacological treatment. We also observed significant sex-specific associations with need for pharmacological treatment for CpG sites within *DRD4* and *AVP*. These findings suggest that epigenetic variation within dopaminergic and stress-response pathways may contribute to variability in NOWS severity among infants with prenatal opioid exposure and that these relationships may differ by infant sex.

We found evidence for epigenetic differences associated with need for pharmacological treatment within *DRD4. DRD4* encodes the dopamine D4 receptor, which mediates dopaminergic signaling pathways implicated in reward processing, behavioral regulation, and feeding behavior. Dopaminergic pathways are known to be altered by opioid exposure and are also implicated in the neurobehavioral manifestations of NOWS.^11^ Genetic studies have linked variation in dopaminergic receptors to substance use disorders and opioid dependence, including polymorphisms in *DRD2* and *DRD4*.^12^ In our study, four CpG sites showed greater DNAm in infants who required treatment relative to those who did not, while one site showed lower DNAm. Notably, multiple CpG sites within the same regulatory region demonstrated consistent associations with treatment status, strengthening confidence that these findings reflect biologically meaningful variation rather than isolated statistical findings. Because increased promoter methylation is generally associated with changes in gene expression, these findings suggest that altered *DRD4* expression could contribute to the behavioral regulation and feeding patterns commonly observed in infants with NOWS.^11^

Although most previous work has focused on genetic variation rather than epigenetic regulation, emerging evidence suggests that *DRD4* methylation may influence behavioral outcomes. Increased methylation of *DRD4* has been associated with greater cognitive and attentional deficits in children with attention-deficit/hyperactivity disorder, indicating that epigenetic regulation of this gene may influence behavioral and regulatory processes.^13^ Prenatal opioid exposure has also been associated with increased risk of attention deficit hyperactivity disorder and related attentional difficulties in childhood.^14^ Given the established role of *DRD4* in attentional regulation and behavioral control, epigenetic variation in this gene may represent one pathway linking NOWS to later neurobehavioral outcomes. While the functional consequences of DNAm in the specific CpG sites examined in our study are unknown, these findings suggest that dopaminergic regulatory mechanisms warrant further investigation in relation to NOWS severity.

We did not find any associations between *OPRM1* methylation and NOWS treatment status after FDR correction. Elevated *OPRM1* methylation has previously been linked to need for pharmacological treatment and NOWS severity, including length of hospital stay and need for multiple medications.^6, 7^ In previous work, we reported that pharmacological treatment for NOWS was associated with decreases in DNAm of *OPRM1* across time, which in turn was associated with improvements in neonatal neurobehavior.^15^ Differences in study design, sample size, tissue source, or the specific CpG sites examined may account for variation across findings.

Despite not replicating earlier associations, our results do suggest that NOWS heterogeneity may arise from dysregulation across dopaminergic and stress pathways, not solely opioid receptor pathways. While prior epigenetic studies of NOWS have focused primarily on *OPRM1*, our findings suggest that broader biological systems involved in reward processing and stress regulation may also contribute to clinically significant NOWS.

To our knowledge, this is the first study to examine sex-specific differences in epigenetic regulation across stress, neurodevelopmental, and dopaminergic pathways in relation to pharmacological treatment for NOWS. Significant Sex × Treatment interactions were detected within *DRD4* and *AVP*, indicating that the relationship between DNAm and need for pharmacological treatment varied by infant sex. Within *DRD4*, multiple CpG sites demonstrated higher DNAm in infants who required pharmacological treatment regardless of sex, suggesting a general association between increased methylation in this regulatory region and need for treatment. However, the CpG site located −362 bp upstream of the transcription start site showed a sex-specific pattern, with higher DNAm only among male infants who required pharmacological treatment and no corresponding difference among females. Together, these findings suggest that different regions within the *DRD4* promoter may reflect both general and sex-specific associations with treatment status. Evidence from developmental research indicates that associations between *DRD4* genotype and behavioral outcomes often differ between males and females, including male-specific associations with novelty-seeking behaviors and stronger associations with ADHD diagnoses in boys.^16, 17^ Some methylation loci may similarly have sex-specific functional implications for NOWS presentation and severity.

The interaction model also found a significant Sex × Treatment interaction within *AVP*, where female infants who required pharmacological treatment exhibited lower DNAm compared to female infants who did not require treatment, although this within-group difference did not reach statistical significance (*p* = .06). *AVP* encodes arginine vasopressin, a neuropeptide involved in hypothalamic stress regulation that may be particularly relevant during prolonged or repeated stress exposure. Sex differences in vasopressin signaling have been reported in stress regulatory brain regions and appear to be influenced by gonadal hormones, suggesting that *AVP* pathways may be differentially regulated in males and females.^18^ Lower DNAm within *AVP* among female infants who required pharmacological treatment may reflect altered regulation of vasopressin signaling involved in stress responsivity, a pattern that has also been observed following prenatal tobacco exposure, supporting the sensitivity of this pathway to in utero substance exposures.^19^ These findings are consistent with prior work demonstrating sex differences in gene expression across dopaminergic and inflammatory pathways in opioid-exposed neonates^5, 20^, supporting the possibility that biological sex shapes how stress and reward pathways respond to prenatal opioid exposure. Sex-specific epigenetic regulation may help explain clinically observed sex differences in NOWS.

Several limitations should be considered when interpreting these findings. Although DNAm was measured prior to initiation of pharmacological treatment, causal inference remains limited, as it is unclear whether observed DNAm differences contribute to withdrawal severity or reflect early physiological processes associated with NOWS. The modest sample size may have limited the ability to detect small effects, particularly after FDR correction. Recruitment from two clinical sites with differing assessment tools and treatment thresholds may introduce residual confounding despite adjustment for study site. Buccal epithelial samples contain mixed cell populations, and we were unable to estimate cell type proportions within the scope of this project, though our approach is consistent with prior candidate gene studies in this population.^6, 7^ Moreover, buccal cells have been proposed as a useful surrogate tissue for DNAm analyses when neural tissue is not accessible.^21^ Our candidate gene approach focused on biologically informed pathways but did not evaluate epigenetic variation across the broader genome. Epigenome-wide approaches may identify additional loci associated with NOWS severity and need for treatment though will require larger samples in order to have adequate power. Finally, without downstream gene expression measurements, the functional consequences of observed DNAm differences in this study can only be hypothesized yet warrant further investigation.

Future research should integrate DNAm and gene expression data with quantitative measures of NOWS severity (e.g., maximum withdrawal scores, duration of pharmacological treatment, use of adjunct medications) as well as newborn and infant neurobehavioral outcomes. Examining these measures together may clarify whether the epigenetic differences identified in this study reflect functional biological pathways contributing to withdrawal severity and neurodevelopment. Larger multisite studies integrating DNAm with gene expression data will be necessary to replicate these findings and clarify their functional significance. Although our findings require replication and functional validation, they suggest that epigenetic variation across dopaminergic and stress-related pathways, alongside sex-specific biological responses to prenatal opioid exposure, may contribute to heterogeneity in NOWS. Identifying reproducible epigenetic markers associated with the need for pharmacological treatment may ultimately support the development of tools for earlier risk stratification among opioid-exposed newborns and improve understanding of the biological pathways underlying NOWS. The convergence of multiple treatment-associated CpG sites within *DRD4* highlights this gene and the broader dopaminergic system as particularly promising targets for future mechanistic investigation.

## Data Availability

All data produced in the present study are available upon reasonable request to the corresponding author, subject to institutional approval and applicable data-use agreements.

## Supplementary

**Table 1:** Maternal and Infant Characteristics by Inclusion Status.

|  | **Overall**<br>**N = 235** | **Included**<br>**N = 181** | **Excluded**<br>**N = 54** |  |
| --- | --- | --- | --- | --- |
|  | M (SD) or N (%) | M (SD)<br>or N (%) | M (SD)<br>or N (%) | **p-value** |
| **Maternal Characteristics** |  |  |  |  |
| *Less than high school degree* | 57 (24%) | 45 (25%) | 12 (23%) | 0.77 |
| *Adequate prenatal care* | 189 (82%) | 146 (81%) | 43 (84%) | 0.75 |
| *No partner* | 107 (46%) | 77 (43%) | 30 (57%) | 0.11 |
| *Hypertension* | 49 (21%) | 39 (22%) | 10 (19%) | 0.80 |
| *Residential Treatment* | 47 (20%) | 40 (22%) | 7 (13%) | 0.20 |
| *Inpatient detox during pregnancy* | 41 (19%) | 34 (20%) | 7 (15%) | 0.60 |
| *Bipolar* | 52 (22%) | 47 (26%) | 5 (9%) | 0.02* |
| *PTSD* | 85 (37%) | 66 (37%) | 19 (36%) | 1.00 |
| *Hepatitis C* | 73 (32%) | 60 (35%) | 13 (25%) | 0.22 |
| **Infant Characteristics** |  |  |  |  |
| *Sex (Male)* | 120 (51%) | 89 (49%) | 31 (57%) | 0.36 |
| *Gestational age (weeks)* | 38.25 (1.71) | 38.29 (1.74) | 38.12 (1.65) | 0.52 |
| *Birth weight (grams)* | 3015.9 (542.6) | 3006.41 (551.08) | 3047.85 (517.03) | 0.61 |
| *Head circumference (cm)* | 33.8 (2.19) | 33.61 (1.88) | 34.27 (2.96) | 0.14 |
| *Birth length (cm)* | 48.4 (3.24) | 48.45 (2.99) | 48.05 (4.03) | 0.52 |
| *Social Services involvement* | 114 (49%) | 85 (47%) | 29 (55%) | 0.42 |
| *Breastfeeding* | 138 (59%) | 108 (60%) | 30 (56%) | 0.70 |
| *Maximum Finnegan Score* | 10.1 (3.01) | 10.3 (3.01) | 9.26 (2.94) | 0.08 |

Note: Percentages are calculated among participants with available data for each variable; denominators may therefore vary across measures. \* $p < 0.05$

## Additional Contributions

We are grateful for the participation of our study infants and their families. We would also like to acknowledge the contributions of James F. Padbury, MD, who enhanced many aspects of our study.

## Abbreviation List

(DNAm): DNA methylation
(NOWS): Neonatal Opioid Withdrawal Syndrome
(FDR): False Discovery Rate
(TSS): Transcription Start Site
(FNAST): Finnegan Neonatal Abstinence Syndrome Scoring Tool

